# Neuroanatomically Derived Genetic Representations Enhance Detection of Plasma Protein Associations

**DOI:** 10.64898/2026.08.19.26360813

**Authors:** Kholod Thaker Alhasani, Upamanyu Ghose, Laura M Winchester, Brian D. Marsden, Cornelia van Duijn, Alejo Nevado-Holgado

**Author notes:** Corresponding author: Alejo Nevado-Holgado.

## Abstract

Genetic variation shapes brain structure, yet it remains unclear whether this neuroanatomical expression of genotype is reflected in circulating proteomic profiles, which provide functional molecular readouts of biological processes. Addressing this question requires integrating genetic, neuroimaging, and proteomic data within the same individuals, which is methodologically non-trivial. To address this challenge, we used brain-genotype scores, an approach recently developed in our lab that allows the creation of individual-level, SNP-specific neuroanatomical representations of genotype learned directly from whole-brain structural MRI. Adapting these scores to proteomics, we tested associations in UK Biobank between 120 brain-genotype scores and plasma levels of 2,920 proteins, followed by pathway and tissue-enrichment analyses to assess biological coherence. After FDR correction, brain-genotype scores yielded 116 significant genomic-neuroimaging-proteomic associations across 49 scores and 52 proteins; none were detected using conventional SNP dosage models, and they explained substantially more variance in protein levels than genotype alone. Enrichment analyses identified convergent immune, metabolic, signalling, and cell-cycle pathways, with tissue enrichment spanning brain, liver, pancreas, and hypothalamus. Several identified proteins overlapped with prior imaging-proteomic literature, supporting biological plausibility. These findings demonstrate that brain-genotype scores reveal biologically meaningful proteomic variation beyond conventional genotype analyses, providing a framework for linking genetic variation, brain structure, and the circulating proteome.

## 1. Introduction

Genetic variation shapes human brain structure across development and ageing, influencing neural architecture, connectivity, and vulnerability to diseases. Large-scale imaging genetic studies have demonstrated that common genetic variants contribute to widespread and coordinated patterns of neuroanatomical variation, rather than isolated effects confined to individual brain regions (Thompson et al. (2020)). At the same time, brain function and pathology are linked to systemic biological processes that can be measured outside the central nervous system. Circulating plasma proteins reflect diverse molecular pathways, including immune regulation, metabolism, synaptic function, and neurovascular signalling, which are often altered in neurodegenerative and neuropsychiatric disorders Dantzer et al. (2008); Heneka et al. (2015); Guest et al. (2015). A key gap is whether genetic effects that are expressed in brain structure are also reflected in circulating protein levels, and whether this can reveal brain-relevant molecular pathways. Addressing this question requires integrating genetic, neuroimaging, and proteomic data within the same individuals.

Imaging genetics, proteomic association studies, and imaging-proteomic studies have largely been conducted independently. Genetic influences on brain structure Elliott et al. (2018); Shen and Thompson (2020); Du et al. (2020), genetic regulation of plasma proteins Sun et al. (2023), and imaging-protein associations Yan et al. (2017); Liu et al. (2025); Ren et al. (2025) are commonly studied using direct associations between two data types at a time. More recently, Ayubcha et al. (2026) used protein quantitative trait loci (pQTLs) from GWAS summary statistics as genetic instruments in a Mendelian randomization framework to estimate causal effects of plasma proteins on brain imaging phenotypes. Multimodal integration methods can combine genetics, neuroimaging, and proteomic measurements by learning shared latent components or cross-modal correlations Du et al. (2021); Zhang et al. (2024). Collectively, these approaches describe global shared variation across modalities or estimate genetically instrumented pairwise effects.

None, however, directly test whether SNP-specific brain patterns relate to circulating proteins, or whether protein associations emerge when genetic information is represented through brain structure rather than genotype alone. The endophenotype concept motivates this approach: biological traits that are closer to gene expression than complex, distal phenotypes can reveal genetic signal not visible in genotype alone Meyer-Lindenberg and Weinberger (2006); Gottesman and Gould (2003). We extend this logic by using brain structure not as an intermediate outcome in a mediation chain, but as a means of generating an enriched, individual-level representation of genotype, which we then test against circulating protein levels.

Here, we use brain-genotype scores that summarise genotype-related patterns in brain structure at the individual level. These scores were developed and validated in prior work using a multi-task 3D CNN trained to predict SNP genotypes from whole-brain T1 MRI Alhasani et al. (2026). We use the resulting probabilistic outputs as continuous measures of how strongly each participant’s brain expresses a SNP-related anatomical pattern. Brain-genotype scores are therefore not the observed genotype itself, but a measure of genotype-related brain patterns that can be tested for association with plasma proteins.

Using UK Biobank genetic, imaging, and plasma proteomics data, we test whether brain-genotype scores are associated with circulating protein levels after adjusting for key genetic and technical factors. We test the hypothesis that these scores capture molecular signatures that extend beyond those detectable using conventional SNP dosage models. To evaluate this, we compare associations identified using brain-genotype scores with conventional SNP–protein tests for the same variants. We then assess the functional coherence of the associated proteins through pathway and tissue enrichment analyses, including brain-relevant resources. Together, this design evaluates whether brain-genotype scores identify protein signatures that are not captured by genotype–protein associations alone, and whether these associations link genotype-related brain patterns to systemic molecular pathways.

## 2. Methods

### 2.1. Derivation of Brain-Genotype Scores

Brain-genotype scores were derived from 3D T1-weighted brain MRI scans and SNP genotype data from the UK Biobank imaging cohort using the framework introduced in our previous work Alhasani et al. (2026). The frame-work learns distributed neuroanatomical representations associated with specific genetic variants and outputs probabilistic genotype scores for each SNP. Brain MRI and genotype data were obtained from the UK Biobank imaging cohort (UKB application 15181). T1-weighted 3D MRI scans (UKB field ID 20252, February 2023 release) underwent the standard UK Biobank preprocessing pipeline, including skull stripping, brain extraction, and nonlinear registration to MNI152 standard space (Alfaro-Almagro et al. (2018)). Analyses were restricted to subjects of White British ancestry (UKB field ID 22006) to minimise population stratification effects.

Genotype data and SNP selection procedures were described previously in Alhasani et al. (2026). In summary, 120 SNPs previously associated with brain T1-MRI image-derived phenotypes (IDPs) were selected after linkage disequilibrium pruning (*r*^2^ < 0.2) and retaining common variants with minor allele frequency (MAF ≥ 0.05). Each SNP genotype was encoded into three classes: 0 (bb, two minor alleles), 1 (Ab, heterozygous), and 2 (AA, two major alleles). The dataset of 41,382 subjects was divided into training (80%), validation (10%), and held-out test (10%) subsets. The testing subset (approximately 4,100 subjects) was held out from all model training, hyperparameter tuning, and architectural decisions. A multi-task 3D CNN was trained to predict SNP genotypes directly from T1-weighted MRI scans. For each SNP, the network produced softmax-normalised probabilities across three genotype classes. These probabilistic outputs constituted the brain-genotype scores used as biomarkers in subsequent proteomic association analyses. In the present study, we used exclusively the CNN-derived brain-genotype scores from the held-out test subset to ensure independence from model training and validation procedures. Detailed descriptions of the CNN architecture, optimisation procedures, and training configuration are provided in Supplementary Methods S1 and in Alhasani et al. (2026). To aid interpretation of brain-genotype scores, gradient-based saliency maps were generated using the explainability framework described in our previous work Alhasani et al. (2026). Saliency maps were used only for visualisation and were not included in the association analyses.

### 2.2. Protein Measurements in Plasma

Proteomic measurements were obtained from UK Biobank participants with available brain-genotype scores in the held-out test subset. Among approximately 4,100 individuals in the CNN test pool, 382 subjects had complete proteomic data and were included in the present analyses (Table 1). Plasma protein concentrations were measured as part of the UK Biobank Pharma Proteomics Project (UKB-PPP) using the Olink Explore 3072 platform, which includes eight panels and quantifies protein levels via proximity extension assay technology (Olink Proteomics AB, Uppsala, Sweden). Sequencing was performed on an Illumina NovaSeq 6000 system (Illumina Inc., San Diego, USA). Olink provides Normalised Protein Expression (NPX) values, which are relative protein abundance measures on a log_2_ scale. NPX values are internally quality controlled and normalised by Olink’s standard pipeline to account for technical variation across plates and batches Sun et al. (2023). Proteins with a missing rate of 30% or higher were excluded (GLIPR1, NPM1, and PCOLCE). After quality control, 2,920 proteins were retained for analysis. Demographic variables (e.g., age and sex) from UKB were used as covariates in the downstream association analyses. All subjects included in this analysis were of White British ancestry, as defined by the UK Biobank genetic ethnicity classification.

**Table 1:**
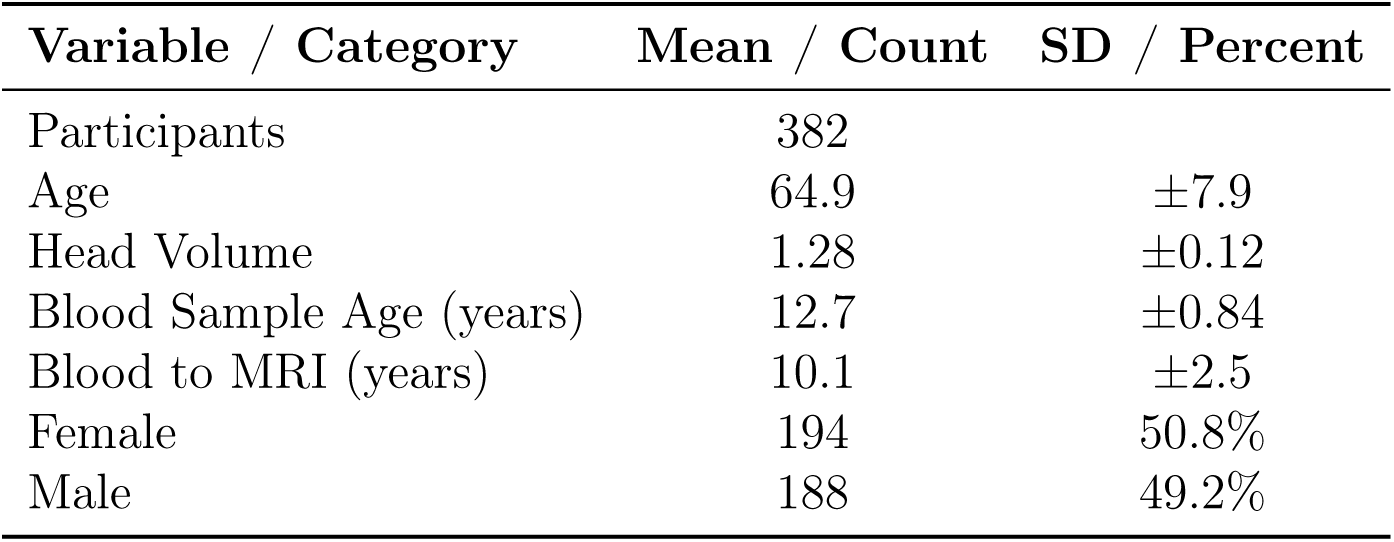
Summary Statistics and Counts.

#### Association Analyses

### 2.3. Association Between Brain-Genotype Scores and NPX

The primary objective of this analysis was to test whether brain-genotype scores were associated with individual differences in NPX values. For each protein, multiple linear regression models were fitted using CNN-derived genotype probabilities (brain-genotype scores) as predictors. To avoid multicollinearity among the three softmax-derived probabilities, only two of the three class probabilities per SNP (corresponding to genotype classes 0, 1, and 2) were included in each model, since the three probabilities sum to one. Covariates included age, sex, top five genetic principal components, genotype array, assessment centre, scanning site, sample age (interval between blood sampling and Olink processing), time between baseline and imaging visit (blood-to-imaging), and Olink batch. For each protein, two nested models were compared: (i) a full model included brain-genotype scores and confounders, and (ii) a reduced model including confounders only. An F-test was used to assess whether inclusion of the brain-genotype scores significantly improved model fit based on residual sum of squares (RSS) differences. Effect sizes were quantified as the change in coefficient of determination (Δ*R*^2^) between the full and reduced models, representing the additional variance in protein expression values explained by the brain-genotype scores beyond that captured by covariates. To examine model robustness, analyses were repeated with alternative confounder adjustments. Specifically, the number of genetic PCs was varied (five and ten PCs) to evaluate whether including additional ancestry components improved model stability given the relatively small sample size. Global head-size scaling was also added as a covariate in sensitivity analyses to test whether adjusting for overall brain volume affected the strength of observed associations. Body mass index (BMI) was additionally included as a covariate in a further sensitivity analysis, given its established association with circulating plasma proteins Sun et al. (2023), to evaluate whether adiposity-related confounding influenced the observed associations. Datasets used in this analysis are summarised in Table 1.

### 2.4. Association of Individual Probability Components with NPX

As an additional validation analysis, we tested each brain-genotype-class probability separately to evaluate the independent contribution of individual probability components to the observed associations. The aim was to evaluate whether using single probability components as predictors would yield a greater, comparable, or reduced number of significant associations relative to the main model that included multiple probability components (brain-genotype scores). In addition, this analysis allowed assessment of the directionality of associations, which cannot be represented by a single regression coefficient in the main multi-component model. For each SNP, three separate models were fitted, each including only one of the CNN-derived probability scores corresponding to genotype classes as the sole predictor. The outcome variable and covariate set were identical to those used in the main association analysis (age, sex, genetic principal components, genotype array, assessment centre, scanning site, sample age, blood-to-imaging interval, and Olink batch). Using the same approach as described for the main analysis, for each protein, nested model comparison was performed between (i) a full model including the single probability score and covariates, and (ii) a reduced model including covariates only. An F-test was used to evaluate whether inclusion of each individual probability score significantly improved model fit based on residual sum of squares (RSS) differences. The change in coefficient of determination Δ*R*^2^ between the full and reduced models was used to quantify the variance in protein expression values explained by each single probability component. Because each model included only a single predictor, standardised regression coefficients (*β*) were extracted to evaluate the direction and magnitude of associations. These coefficients were used to construct volcano plots displaying (*β*) versus statistical significance, providing a descriptive assessment of effect directionality. This analysis was conducted as a validation step to characterise the behaviour of individual probability components and to compare their performance with the main brain-genotype score model.

### 2.5. Comparison of Brain-Genotype Scores and Actual Genotypes

To compare CNN-derived brain-genotype scores with direct genetic effects, additional association analyses were performed using raw SNP genotypes as predictors. Each SNP was modelled under an additive framework, coded as a dosage variable (0, 1, 2) representing the number of minor alleles. For each protein, linear regression models were fitted with genotype dosage as the main predictor and the same covariates used in the brain-genotype score analyses. Using the same approach as described for the main analysis, two nested models were compared: (i) a full model including genotype dosage and covariates, and (ii) a reduced model including covariates only. An F-test was used to determine whether inclusion of genotype dosage significantly improved model fit based on residual sum of squares (RSS) differences. Effect sizes were quantified as the change in coefficient of determination (Δ*R*^2^) between the full and reduced models, representing the proportion of additional variance in protein expression values explained by genotype dosage. Results from these models were compared with those obtained using brain-genotype scores to evaluate whether the CNN-derived scores captured similar or distinct proteomic associations relative to direct genotypic variation.

As an additional sensitivity analysis to enable a degrees-of-freedom-matched comparison between the two modelling approaches, we further derived a single expected brain-genotype dosage per SNP, calculated as the probability-weighted sum of genotype classes (0×P0 + 1×P1 + 2×P2) from the CNN-derived brain-genotype scores, analogous to expected dosage calculations used in genotype imputation. This single continuous score was tested as the sole predictor of NPX using the same nested-model regression framework described above, allowing direct comparison with the actual genotype dosage model under an identical single-predictor structure.

### 2.6. Functional and Biological Annotation

Functional and biological annotation analyses were performed for proteins significantly associated with brain-genotype scores, including pathway enrichment and chromosomal distribution analyses. Functional enrichment was conducted using the gseapy Python package, which provides access to Enrichr gene set libraries. For each SNP, the set of proteins significantly associated with its brain-genotype score (raw *p* <= 0.05) was used as input. The background gene set comprised all 2,920 protein-coding genes quantified in the UK Biobank Olink dataset, ensuring enrichment was evaluated relative to the available proteomic coverage. Analyses were performed separately for the KEGG, Reactome, GTEx, and Allen Brain Atlas libraries. To reduce redundancy among enriched terms, enrichment results within each gene-set library were collapsed into broader functional categories using the hierarchy files provided by the respective databases. For each SNP, raw p-values of terms belonging to the same higher-level category were combined using the Aggregated Cauchy Association Test (ACAT) to obtain a single aggregated p-value per category. Multiple testing correction was then applied within each gene set library across all SNPs using the Benjamini–Hochberg false discovery rate (FDR) procedure. Pathways with FDR-adjusted *p* < 0.05 were considered significantly enriched. For chromosomal distribution analysis, significantly associated protein-coding genes were mapped to chromosomes using HGNC gene symbol identifiers. For each chromosome, the number of significant genes was divided by the chromosome length to calculate the density of associated proteins, accounting for differences in chromosome size.

## 3. Results

### 3.1. Plasma Proteins Associated with SNP Brain-Genotype Scores

We first investigated the associations between 2,920 plasma proteins and 120 brain-genotype scores, each corresponding to a single SNP. Overall, 116 significant associations were identified between 49 brain-genotype scores and 52 proteins after the false discovery rate (FDR) correction for multiple comparisons (FDR < 0.05). The proteins involved in the greatest number of significant associations were NFIC, DLGAP5, IGFL4, BTLA, and RASGRF1. The brain-genotype scores with the largest numbers of significant associations corresponded to SNPs rs3922583, rs17010085, rs1874581, rs13161403, and rs11762352.

The ten most significant associations are summarised in Table 2, including the full model *R*^2^, *p*-values, and corresponding SNP identifiers. Detailed results for all tested brain-genotype score–protein pairs are provided in Supplementary material. Figure 1 A illustrates the overall relationship between effect sizes (Δ*R*^2^) and statistical significance (− log_10_ *p*-values) across the UKB proteomic panel. To further characterise the structure of the significant associations, hierarchical clustering was performed on all FDR-significant brain-genotype scores–protein pairs and visualised as a clustered heatmap (Figure 2). To illustrate the neuroanatomical information encoded by brain-genotype scores, we visualised saliency maps for three representative scores among those showing the strongest protein associations (Figure 3). These maps illustrate the distributed neuroanatomical patterns contributing to each score, supporting their interpretation as biologically meaningful neuroanatomical representations. Sensitivity analyses were performed by additionally adjusting for global head volume scaling factor, BMI, and additional genetic principal components. Most significant brain-genotype score–protein associations were robust across sensitivity models, with 72 associations remaining FDR-significant in all analyses with substantial overlap observed among the top associations (Figure 4). Adjustment for head volume produced the largest change in the set of significant associations, whereas BMI adjustment and inclusion of additional principal components had comparatively minor effects. Effect sizes (Δ*R*^2^) were largely stable across models. Detailed results are provided in Supplementary S2 Figure 1, and Supplementary Table.

**Figure 1:**
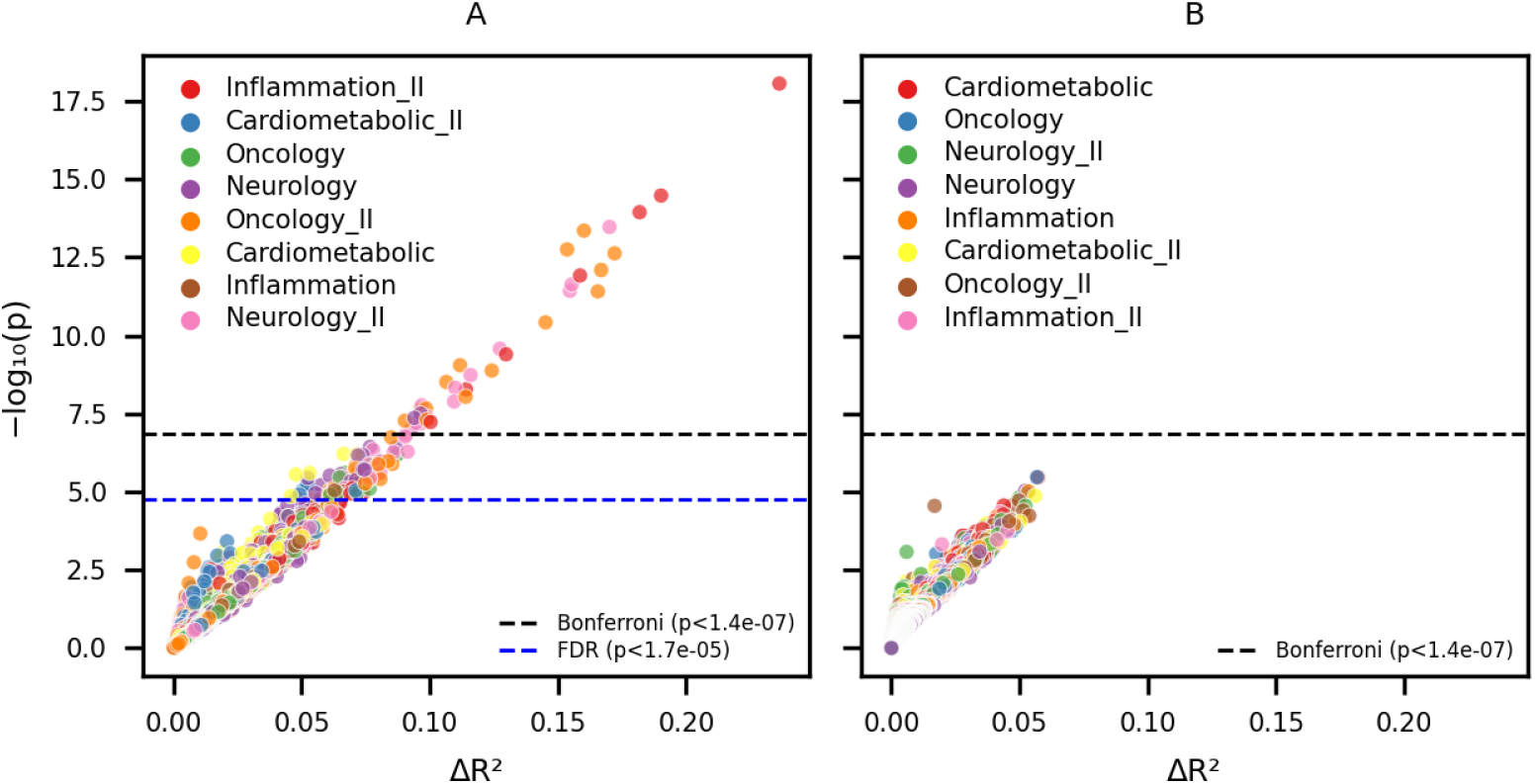
Association results for brain-genotype score and direct genotype models. Scatter plots showing statistical significance (− log_10_(*p*), y-axis) versus additional variance explained (Δ*R*^2^, x-axis) for protein associations. Each point represents one SNP–protein association tested using the same linear regression framework and covariate set. (A) Results obtained using brain-genotype scores as predictors. (B) Result when using observed SNP dosage values (0, 1, 2) as predictors. The black dashed line indicates the Bonferroni significance threshold. The blue dashed line indicates the FDR threshold ((q < 0.05)) and appears only in (A), as no associations remained FDR-significant in the direct genotype model.

**Figure 2:**
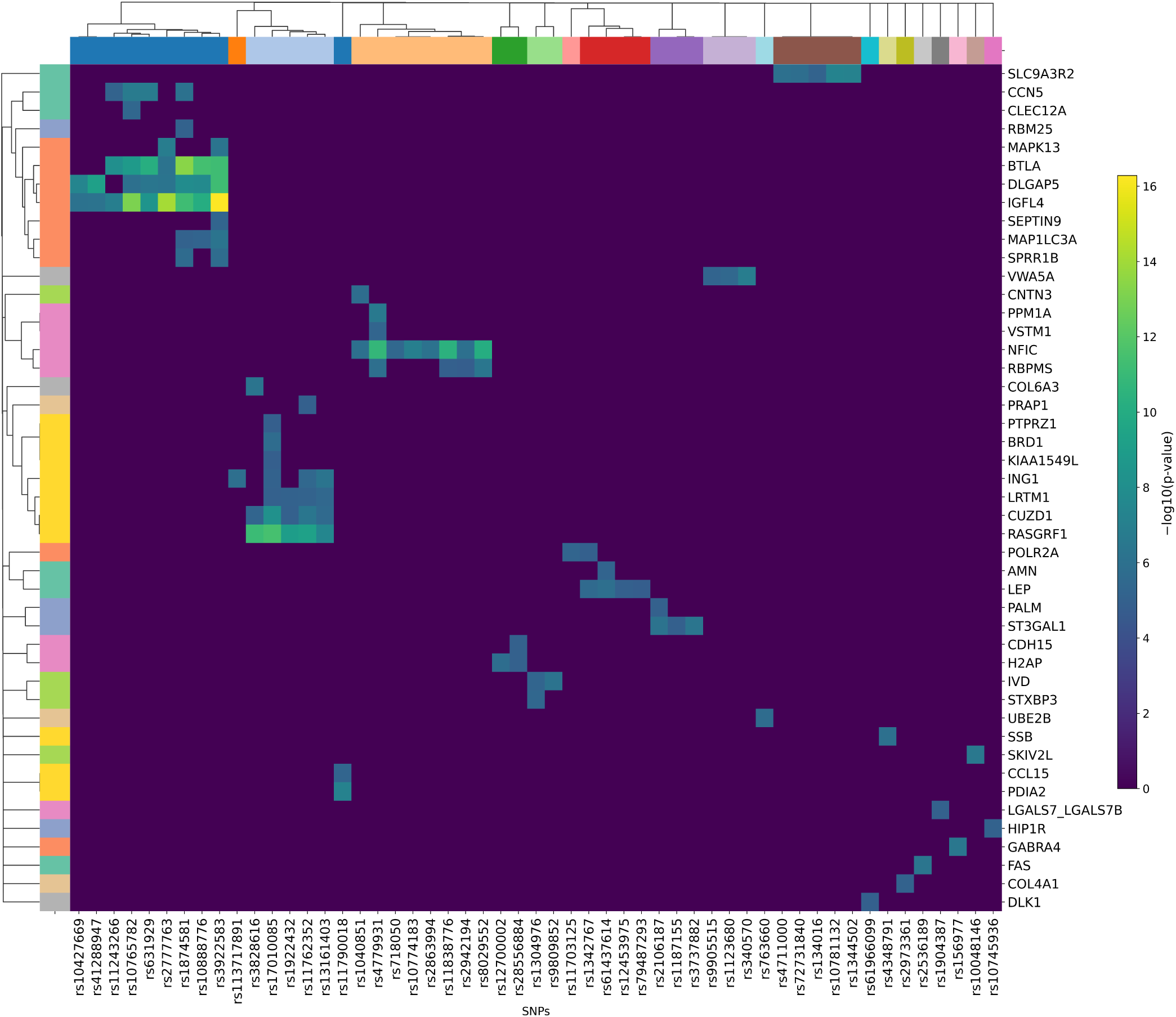
Hierarchical clustered heatmap of FDR-significant associations between brain-genotype scores and plasma proteins. Rows represent plasma proteins and columns represent brain-genotype scores (SNPs). Cell colour indicates association strength (log10(FDR-adjusted p-value)). Hierarchical clustering was applied to both rows and columns to group proteins and brain-genotype scores with similar association patterns. Only FDR-significant associations are shown.

**Figure 3:**
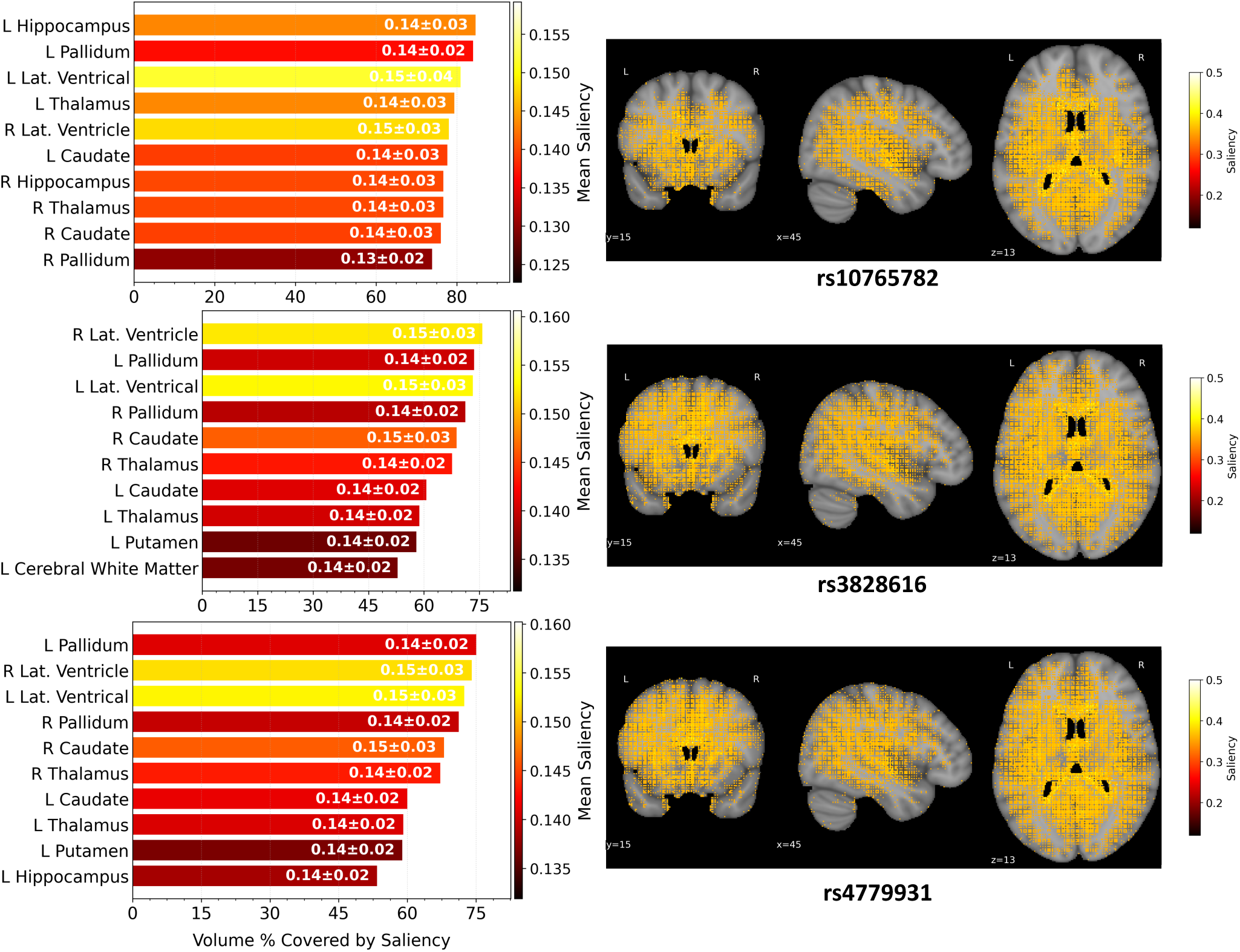
Anatomical interpretation of representative brain-genotype scores. Saliency maps for three representative brain-genotype scores (rs10765782, rs3828616, and rs4779931) showing the brain regions contributing most strongly to the CNN predictions. For each SNP, the left panel shows the ten anatomical regions with the greatest saliency contribution, expressed as the percentage of regional volume covered by salient voxels together with the mean saliency intensity (mean ± SD). The right panel displays the corresponding whole-brain saliency maps overlaid on coronal, sagittal, and axial MRI views.

**Figure 4:**
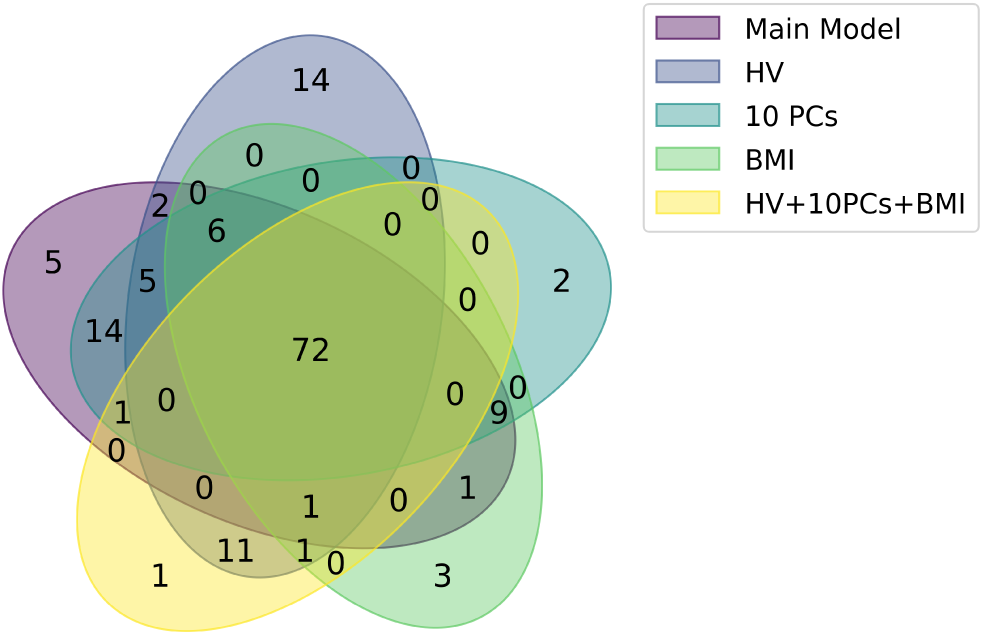
Overlap of FDR-significant brain-genotype score–protein associations across sensitivity analyses. Venn diagram comparing associations identified in the main model, head volume-adjusted model (HV), 10 principal components-adjusted model (10 PCs), BMI-adjusted model, and the combined model (HV + 10 PCs + BMI). Numbers indicate associations unique to or shared among models.

**Table 2:** Summary of the top 10 SNP-score associations with corresponding proteins, *R*^2^ values, and FDR-adjusted *p*-values. Full results are available in Supplementary Table S1.

| SNP rsid | Protein | n samples | $R^2$full model | Panel | Chr | p-value |
| --- | --- | --- | --- | --- | --- | --- |
| rs3922583 | IGFL4 | 304 | 0.3507 | Inflammation | 19 | $3.00 \times 10^{-13}$ |
| rs2777763 | IGFL4 | 311 | 0.3068 | Inflammation | 19 | $5.79 \times 10^{-10}$ |
| rs10765782 | IGFL4 | 316 | 0.2947 | Inflammation | 19 | $1.32 \times 10^{-9}$ |
| rs4779931 | NFIC | 346 | 0.2296 | Neurology | 19 | $2.93 \times 10^{-9}$ |
| rs3828616 | RASGRF1 | 362 | 0.2262 | Oncology | 15 | $3.09 \times 10^{-9}$ |
| rs17010085 | RASGRF1 | 362 | 0.2196 | Oncology | 15 | $1.03 \times 10^{-8}$ |
| rs1874581 | BTLA | 314 | 0.2605 | Oncology | 3 | $1.17 \times 10^{-8}$ |
| rs10888776 | BTLA | 310 | 0.2602 | Oncology | 3 | $3.47 \times 10^{-8}$ |
| rs1874581 | IGFL4 | 315 | 0.2710 | Inflammation | 19 | $4.59 \times 10^{-8}$ |
| rs3922583 | DLGAP5 | 312 | 0.2755 | Neurology | 14 | $7.99 \times 10^{-8}$ |

### 3.2. Comparison of Brain-Genotype Scores and Actual Genotypes

When testing the direct genotype dosage of each SNP against protein expression levels, no SNP–protein associations reached statistical significance after false discovery rate (FDR) correction (Figure 1B). In contrast to the main model using brain-genotype scores (Figure 1A), the distribution of effect sizes was markedly reduced. Although nominal associations were observed, none exceeded multiple-testing thresholds.

To directly match the number of predictors used in the genotype dosage model, we repeated the primary analysis using a single expected brain-genotype dosage per SNP instead of the two-parameter probability representation. This single-predictor brain-derived model identified 28 significant SNP–protein associations after FDR correction, involving 19 unique brain-genotype scores and 16 proteins (Supplementary S2 Figure 3, Supplementary results), compared with zero significant associations for the actual genotype dosage model. Effect sizes remained comparable in magnitude to the primary two-parameter model, with Δ*R*^2^ values reaching up to 0.23. The strongest associations again involved IGFL4, NFIC, BTLA, and DLGAP5, consistent with the top signals identified in the primary analysis (Table 2). The number of significant associations was lower than in the primary two-parameter model (28 vs. 116).

### 3.3. Functional and Biological Annotation of Brain-Genotype-Associated Proteins

Functional and biological enrichment analyses were performed for each SNP to assess whether the associated proteins were enriched in specific biological or functional processes (Figure 5). KEGG and Reactome gene sets identified consistent enrichment of immune, metabolic, and signalling pathways across independent annotation systems (Figure 5 A,C). In KEGG, the most frequently enriched categories included immune system pathways (7 SNPs, minimum FDR-adjusted *p* = 1.22 × 10*^−^*^5^), metabolism (7 SNPs, *p* = 2.81 × 10*^−^*^5^), digestive system pathways (6 SNPs, *p* = 2.35 × 10*^−^*^5^), and signal transduction pathways (3 SNPs, *p* = 1.32 × 10*^−^*^4^). Reactome showed a similar pattern, with metabolism enriched for 7 SNPs (minimum FDR-adjusted *p* = 4.21 × 10*^−^*^5^), signal transduction (4 SNPs, *p* = 5.76 × 10*^−^*^7^), and immune-related pathways (3 SNPs, *p* = 2.08 × 10*^−^*^6^). Several SNPs, including rs10888776, rs7589256, and rs61966099, showed overlapping enrichment across KEGG and Reactome categories. GTEx tissue enrichment analysis showed that associated genes were most frequently enriched in brain (10 SNPs, minimum FDR-adjusted *p* = 1.39 × 10*^−^*^5^), pancreas (7 SNPs, *p* = 4.99 × 10*^−^*^6^), and liver tissues (4 SNPs, *p* = 2.54 × 10*^−^*^7^), reflecting both neural and metabolic contexts for the identified associations (Figure 5B). Allen Human Brain Atlas revealed region-specific expression patterns associated with individual brain-genotype scores (Figure 5E). The broadest regional enrichments were observed for rs10888776 and rs61966099, which were enriched across 11 and 5 brain regions, respectively. Across SNPs, the hypothalamus (6 SNPs, minimum FDR-adjusted *p* = 4.55 × 10*^−^*^9^) and frontal cortex (5 SNPs, *p* = 7.26 × 10*^−^*^5^) were the most frequently enriched regions. For chromosomal distribution, after accounting for chromosome length, chromosome 19 showed the highest density of significantly associated proteins, followed by chromosomes 20, 14, 16, and 15 Figure 5 panel D.

**Figure 5:**
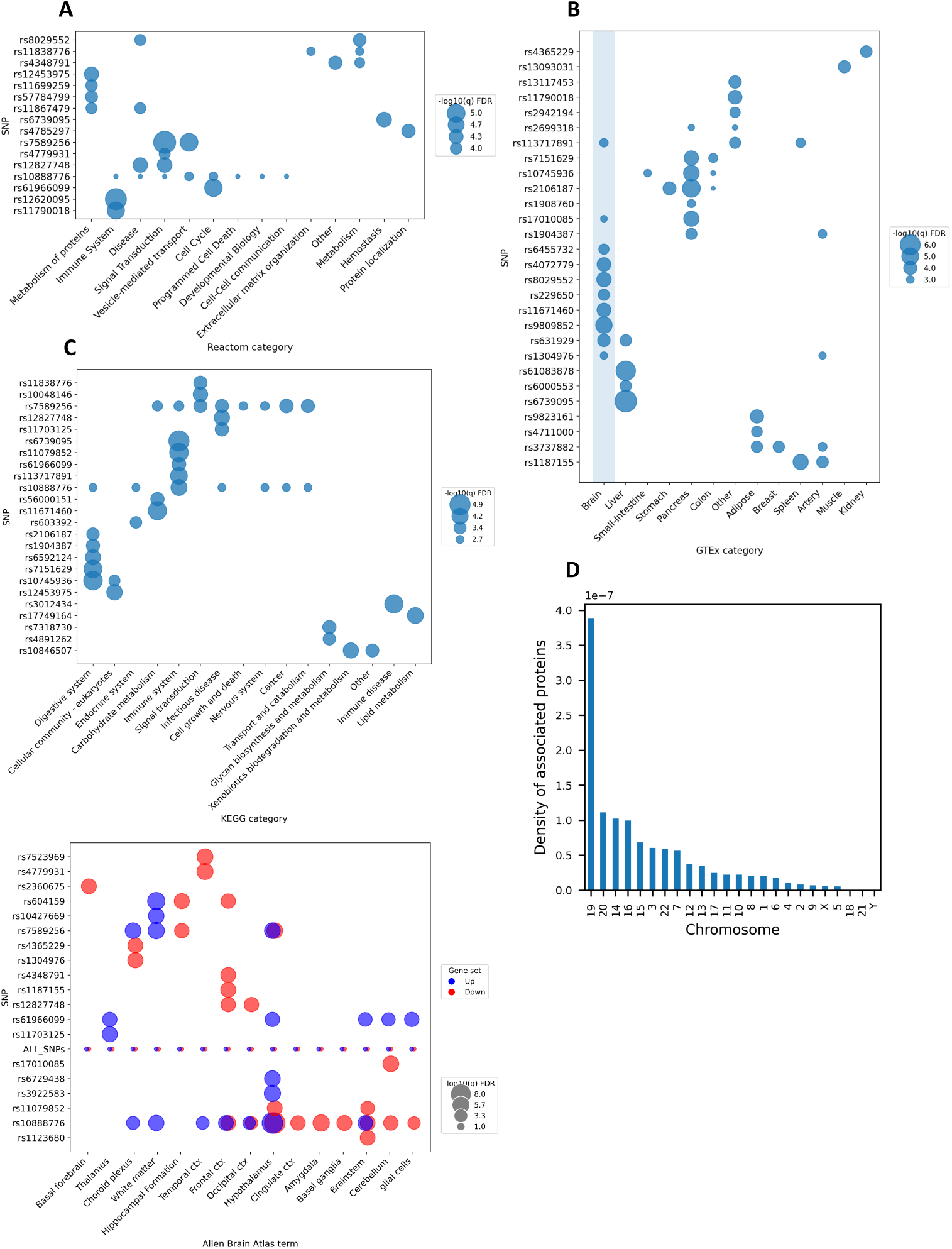
Functional enrichment of proteins associated with brain–genotype scores. (A) KEGG pathway enrichment. (B) GTEx tissue enrichment. (C) Reactome pathway enrichment. (D) Chromosomal distribution of associated protein-coding genes normalised by chromosome length. (E) Allen Human Brain Atlas regional enrichment; bubble colour indicates gene expression direction. In panels A, B, C, and E, bubble size is proportional to − log_10_(FDR).

### 3.4. Association of Individual Probability Components with NPX and Directionality Analysis

To evaluate whether the joint brain-genotype score provides additional information beyond its individual components, each genotype-class probability (*P*_0_, *P*_1_, and *P*_2_), corresponding to the CNN-estimated probabilities of genotype classes 0, 1, and 2, was tested separately as a predictor of NPX. Compared with the main model using combined brain-genotype scores, the single-component models identified fewer significant SNP–protein associations. After FDR correction, the individual probability models identified 36 (*P*_0_), 58 (*P*_1_), and 37 (*P*_2_) significant SNP–protein pairs, compared with 116 significant pairs in the main model. The number of unique SNPs and proteins reaching significance was also reduced in each individual component analysis. The range of explained variance (Δ*R*^2^) was smaller for all three probability components than for the main model, with lower maximum Δ*R*^2^ values observed across all single-component analyses (Supplementary S3 Figure 2).

As the brain-genotype score is evaluated jointly through multiple probability components, its overall effect cannot be summarised by a single regression coefficient. Therefore, the individual probability models were additionally used to examine the direction of associations. Standardised regression coefficients (*β*) were visualised using volcano plots (Figure 6). Probability classes 0 and 1 showed similar distributions of effect sizes, with significant associations distributed on both positive and negative sides of *β* = 0. Probability class 2 showed a comparable overall pattern but fewer positive associations and a narrower effect-size range. Together, these analyses indicate that modelling the genotype-class probabilities jointly captures more associations than analysing each probability separately, while the individual probability models provide an interpretable view of association directionality.

**Figure 6:**
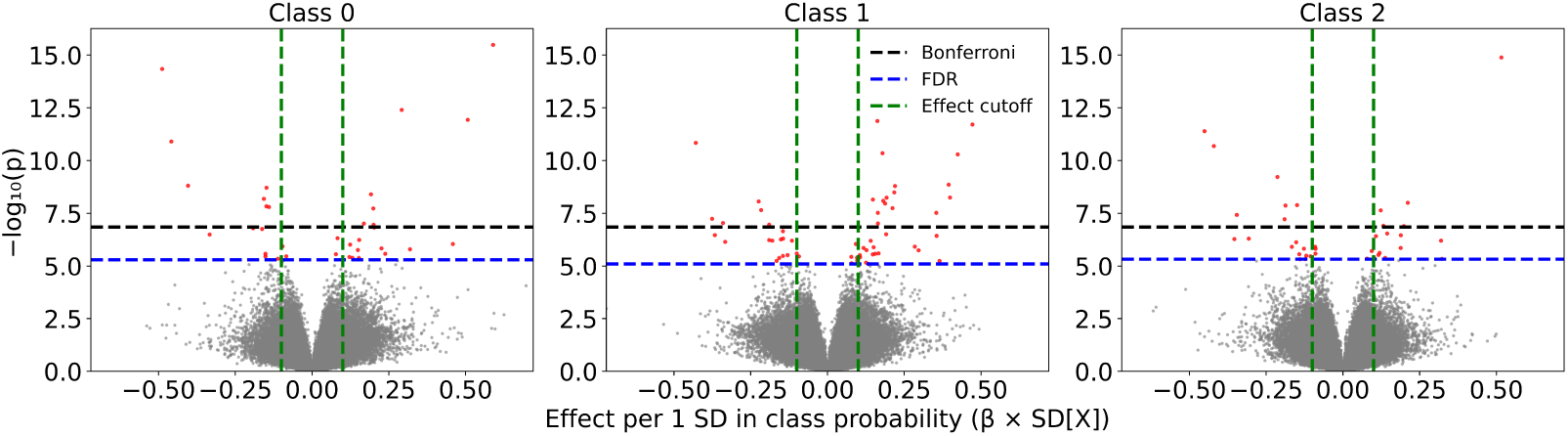
Volcano plots for the individual probability components of the brain-genotype scores. Volcano plots showing the standardized regression coefficient (*β* per 1 SD increase in predictor; x-axis) against statistical significance (− log_10_(*p*); y-axis) for associations between plasma proteins and each of the three CNN-derived genotype-class probability components (P0, P1, and P2). Each point represents a single SNP–protein association tested using the same regression framework as the primary analysis. Red points indicate associations remaining significant after FDR correction.

## 4. Discussion

In this study, we identified significant associations between brain-genotype scores and circulating plasma proteins, revealing molecular correlates of genetically influenced brain structure. In total, 116 associations were observed across 49 unique SNP-based scores and 52 proteins, with the strongest signals involving IGFL4, NFIC, RASGRF1, BTLA, and DLGAP5. These associations were not detected using conventional SNP–protein models based on additive genotype coding. Moreover, the probabilistic brain-derived representation explained a greater proportion of variance in protein expression and yielded more significant associations than models using either raw genotype dosage or individual genotype-probability components. Together, these findings indicate that modelling genetic variation through its expression in neuroanatomical structure provides improved sensitivity for detecting molecular associations.

Pathway enrichment analyses of proteins associated with individual brain-genotype SNP scores revealed convergent biological themes. The most consistently enriched categories involved immune processes, metabolic and digestive pathways, signal transduction, and cell-cycle regulation. These domains extend beyond neuron-specific mechanisms and suggest that genetic influences on brain structure intersect with broader physiological systems. Tissue enrichment further highlighted expression profiles in brain, liver, and pancreas, aligning with the metabolic and immune themes identified at the functional level. Regional enrichment in the hypothalamus, an integrative centre for endocrine and metabolic regulation Tran et al. (2022), adds anatomical plausibility to these findings. Although enrichment analyses do not establish causality, the coherence of functional and tissue-level patterns supports the presence of shared biological substrates linking neuroanatomy and peripheral molecular regulation. The observed associations are consistent with genetic pleiotropy, whereby common variants exert effects across multiple tissues and physiological systems. Because brain-genotype scores summarise genetic variation expressed through structural brain phenotypes, their relationships with circulating proteins suggest that part of the genetic architecture shaping neuroanatomy also contributes to systemic molecular traits. This interpretation aligns with prior literature suggesting that blood-based biomarkers reflect bidirectional interactions between peripheral physiology and the brain Chan et al. (2014); Guest et al. (2015), and supports a view of structural brain variation as embedded within broader regulatory networks rather than isolated neural processes.

Recent imaging–proteomic studies in the UK Biobank provide additional context for our findings. Liu et al. (2025) reported associations between brain-age gap scores derived from imaging and plasma proteins, identifying 13 biomarkers including BCAN. In our analysis, BCAN was also significantly associated with two SNP-based brain-genotype scores (rs11762352 and rs6592124), suggesting convergent biological signals. Similarly, Ren et al. (2025) identified 1,143 proteins associated with brain imaging measures, of which 21 overlapped with proteins identified in our study (including NFIC, CCN5, SLC9A3R2, MAP1LC3A, and ING1). A complementary analysis by Ayubcha et al. (2026), using Mendelian randomization to combine protein quantitative trait loci from UKB-PPP and deCODE with UK Biobank imaging phenotypes, identified colocalization-supported associations across 28 proteins, including BCAN (linked to white-matter tract microstructure) and NTF3, both of which were also significant in our study. BCAN has thus now been identified independently across three separate analyses, including our own, while NTF3 represents a further overlap unique to this comparison. Although each study used a different analytic approach — direct phenotypic correlation, genetic instrumentation, or our own imaging-genotype-to-protein design — none tested the same genotype–protein relationship examined here. The recurring overlap, especially for BCAN, therefore reflects convergent evidence across independent frameworks rather than direct replication, strengthening the biological plausibility of our findings and suggesting that part of the imaging–proteomic relationship may reflect shared underlying genetic architecture.

Beyond biological interpretation, this study also demonstrates a methodological contribution in modelling genetic variation through neuroanatomical structure. Rather than relying solely on conventional additive SNP coding, we used brain-derived probabilistic representations of genotype as predictors in protein association analyses and directly compared their performance with standard genotype models. The brain-informed representation yielded a greater number of detectable associations and explained more variance in protein expression, indicating that conditioning genetic information on its neuroanatomical expression enhances sensitivity to downstream molecular effects. Because brain-genotype scores are derived jointly from genotype labels and whole-brain structural MRI, they incorporate a richer, higher-dimensional source of individual-level variation than a single dosage value, which may further explain their improved detection power relative to conventional genotype coding. This framework addresses a broader challenge in multi-omic research, where individual SNP effects are often small and different biological modalities are analysed independently. By transforming static genotype coding into a biologically informed representation grounded in brain structure, this approach provides a complementary strategy for integrating genetic variation with systemic phenotypes. Given that similar representations have been applied to other outcomes, such as cognitive traits in prior work, this modelling strategy may be generalisable to additional molecular or clinical phenotypes.

### 4.1. Limitations and Future Directions

Several limitations should be considered. First, the analysis was restricted to 120 SNP-based brain-genotype scores derived from variants with established associations with structural brain measures, and to a relatively small sample of 382 individuals with paired imaging and proteomic data. By comparison, large-scale pQTL studies such as Sun et al. (2023) achieve genome-wide significance using approximately 54,000 participants and an unbiased, genome-wide variant set; the absence of significant associations in our direct genotype model may therefore partly reflect limited statistical power and the restricted, non-random SNP panel, rather than solely the superiority of the brain-derived representation. While this targeted approach ensured biological relevance, it does not capture genome-wide variation and may overlook additional loci contributing to brain–protein relationships. Extending this framework to a broader set of SNPs, including genome-wide representations, may provide a more comprehensive view of genetically mediated brain–body interactions.

Second, the study is based on cross-sectional data, limiting inference regarding temporal relationships among genetic variation, brain structure, and protein expression. Although the observed associations are consistent with shared genetic architecture, they do not establish mediation or directionality. Future studies incorporating longitudinal data, formal mediation modelling, or Mendelian randomisation approaches could help clarify potential causal pathways. Future research may extend this framework beyond plasma proteomics to other molecular domains, such as metabolomics or epigenetic data, to determine whether brain-informed genetic representations consistently enhance cross-modal integration. Applying this approach in independent cohorts and disease-specific samples will further clarify its robustness and potential translational relevance.

## 5. Conclusions

In this study, we demonstrate that brain-genotype scores, probabilistic representations of genetic variation derived from structural MRI, are significantly associated with circulating plasma protein expression. Compared to conventional SNP dosage models, these brain-informed genetic representations improved detection of molecular associations and explained greater variance in protein levels. Enrichment analyses revealed convergent immune, metabolic, and signalling pathways, alongside tissue and regional patterns consistent with coordinated brain–body regulation. Together, these findings support a framework in which genetic influences on brain structure represent one expression of broader molecular networks spanning central and peripheral systems.

## Data Availability

All data produced in the present study are available upon reasonable request to the authors

## Author contributions

KTA conceptualised the study, developed the analysis code, performed the analyses, and wrote the manuscript. ANH provided primary supervision, contributed to study design and interpretation of results. LW, BM, CVD provided ongoing supervision, contributed to the interpretation of the findings. All authors contributed to the drafting and revision of the manuscript for intellectual content. All authors have read and agreed to the published version of the manuscript.

## Data availability

UK Biobank data is available via application on the cohort website. https://www.ukbiobank.ac.uk/enable-your-research/apply-for-access

## Acknowledgments

This research has been conducted using the UK Biobank Resource under Application Number 15181.

