## Supplementary_Material for "Neuroanatomically Derived Genetic Representations Enhance Detection of Plasma Protein Associations"

Supplementary material for:
*Neuroanatomically Derived Genetic Representations Enhance Detection of Plasma Protein Associations*

### 1. CNN model and Derivation of Brain-Genotype Scores

A lightweight 3D convolutional neural network (CNN) was developed to derive brain-genotype scores from T1-weighted MRI imaging. The model architecture followed the design introduced in our previous work Alhasani et al. (2026), which adopted a ResNet-10 backbone Kaiming He and et al. (2015) for volumetric data by replacing standard 2D layers with their 3D equivalents. Model inputs were MRI volumes with dimensions of 182 x 218 x 182 voxels and 1 mm isotropic resolution. The CNN model was trained as a multi-task classifier that simultaneously predicted genotype probabilities for multiple SNPs per subject. Each SNP was treated as an independent prediction head with three genotype classes: 0 (minor-allele homozygote), 1 (heterozygote), and 2 (major-allele homozygote). This setup allows the network to learn shared anatomical representations across SNPs while producing SNP-specific probabilistic outputs interpreted as brain-genotype scores. To balance computational efficiency and convergence stability, the final configuration jointly predicted 10 SNPs per model instance. Each network therefore produced an output tensor of size $T \times 3$ ($T = 10$) , where each row contains the softmax-normalised class probabilities for a given SNP. Twelve such models were trained to cover all 120 variants included in the analysis. The CNN architecture starts with an initial 7×7×7 convolution followed by batch normalisation Ioffe and Szegedy (2015), ReLU activation and max pooling **?**. Four residual blocks followed, each integrating a Squeeze-and-Excitation (SE) module Hu et al. (2017) to enhance channel-wise feature calibration. (Equation 1).

$$\mathbf{z}_c = \frac{1}{HWD}\sum_{i,j,k}\mathbf{X}_{c,i,j,k}, \qquad \mathbf{s} = \sigma(\mathbf{W}_2\,\delta(\mathbf{W}_1\mathbf{z})), \qquad \tilde{\mathbf{X}}_c = \mathbf{s}_c\mathbf{X}_c. \tag{1}$$

Global average pooling generated a 512-dimensional representation per subject which was passed to SNP-specific fully connected layers to yield the genotype probability distributions. Training was performed in PyTorch using the AdamW optimiser with multi-task cross-entropy objective (per-task losses summed). The hyperparameters learning rate, batch size, and weight decay were tuned with Optuna across 50 trials. The best configuration used a learning rate of $5 \times 10^{-6}$, batch size 20, and weight decay $8.01 \times 10^{-5}$. Equal task weights were maintained to avoid bias towards specific SNPs. The resulting softmax probabilities for each SNP served as quantitative neuroanatomical scores used in all subsequent association analyses with normalised protein expression. An overview of the model architecture is illustrated in Figure **??**.

### 2. Association Between Brain-Genotype Scores and Normalised Protein Expression: Sensitivity Models

To evaluate the robustness of the main association results, additional regression models were fitted using the same statistical framework as the primary analysis Figure 1. 1- Head-volume–adjusted model: Global head volume (intracranial scaling factor) was added as an additional covariate to assess whether adjusting for overall brain size influenced the observed associations. 2- Extended ancestry model (10 PCs): The number of genetic principal components was increased from five (used in the main model) to ten, to examine whether inclusion of additional ancestry components affected association results. 3- Body mass index (BMI)–adjusted model: Given its established association with circulating levels of plasma proteins, to evaluate whether adiposity-related confounding influenced the observed associations. 4- All additional confounds together. All other covariates (age, sex, genotype array, assessment centre, scanning site, sample age, blood-to-imaging interval, and Olink batch) were identical to those used in the primary analysis. Nested model comparison and FDR correction procedures were unchanged.

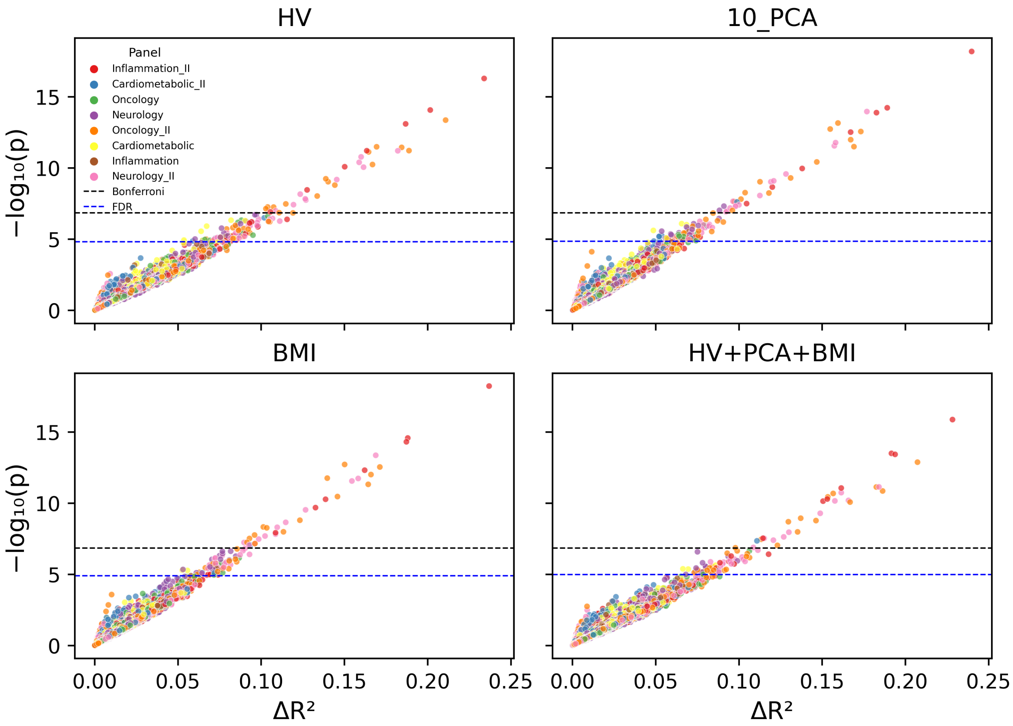


Figure 1: Sensitivity Models

### 3. Association of Single Probability Components of Brain-Genotype Scores

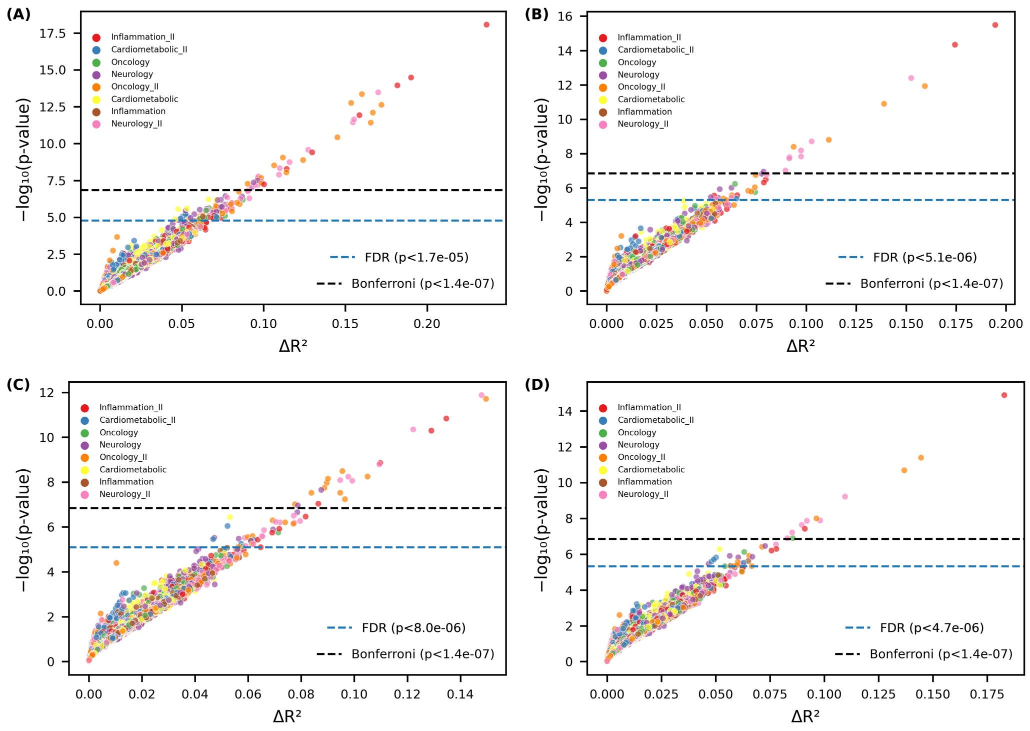


Figure 2: Scatter plots of association results for the main model (combined score) and single-component models. Scatter plots showing statistical significance ($-\log_{10}(p)$) versus additional variance explained ($\Delta R^2$) for associations between brain-genotype scores or individual probability components and normalised protein expression values. Each point represents one SNP–protein model tested using the same linear regression framework and covariate set. (A) Main model using brain-genotype scores (derived from two probability components as predictors). (B) Model using the probability of genotype class 0 (two copies of the minor allele) as predictor. (C) Model using the probability of genotype class 1 (heterozygous). (D) Model using the probability of genotype class 2 (two copies of the major allele). The black dashed line indicates the Bonferroni significance threshold. The blue dashed line indicates the FDR threshold ($q < 0.05$)

### 4. Association Between Actual Genotypes and NPX, and Comparison with Expected Brain-Genotype Dosage

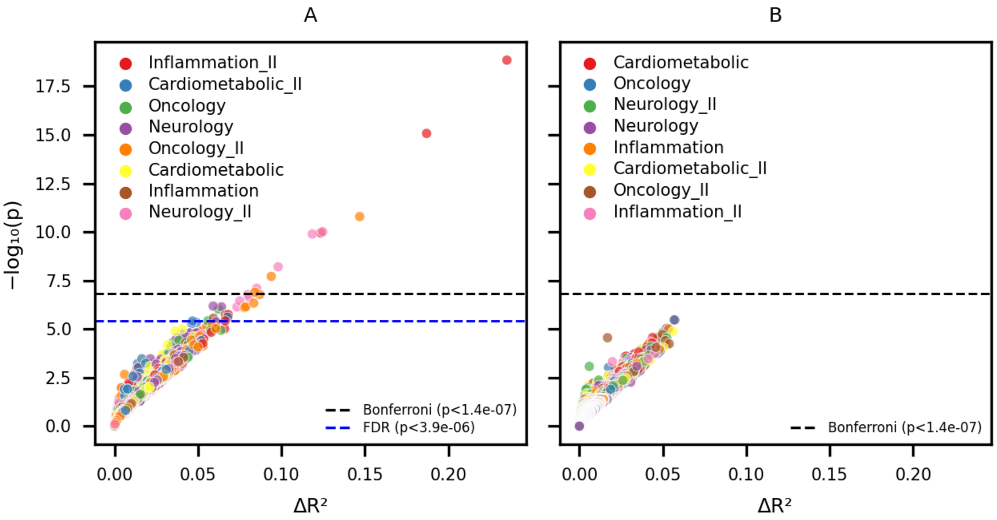


Figure 3: Association results using expected brain-genotype dosage compared with actual genotype dosage. (A) Results obtained using expected brain-genotype dosage as predictors. (B) Results using observed SNP dosage values (0, 1, 2) as predictors. The black dashed line indicates the Bonferroni significance threshold. The blue dashed line indicates the FDR threshold ($q < 0.05$) and appears only in (A), as no associations remained FDR-significant using actual genotype dosage.
